# Dynamic whole-body fetal sensorimotor mapping with three-dimensional cine MRI

**DOI:** 10.64898/2026.11.26.26369024

**Authors:** Hoshinori Kanazawa, Tatsuhiro Wada, Seiichi Morokuma, Yukiyo Shimada, Hiroha Tokui, Koji Sagiyama, Chiaki Tokunaga, Kei Nishikawa, Yuya Saito, Yasuo Kuniyoshi

**Author notes:** These authors contributed equally to this work (Hoshinori Kanazawa, Tatsuhiro Wada).

## Abstract

Early spontaneous whole-body movements generate sensory feedback believed to contribute to sensorimotor maturation, and assessing these movements informs clinical evaluation of neurological development in infants. Yet the sensorimotor conditions of spontaneous fetal movement remain difficult to characterise across the body in vivo. Here we introduce cine MRI–based fetal sensorimotor mapping (cine-FSM), which combines rapid volumetric imaging (2.3–3.3 volumes s^−1^) with body-surface and musculoskeletal modelling. In 11 third-trimester pregnancies, cine-FSM reconstructed whole-body movement and estimated skin contact and muscle lengths. The resulting maps separately captured uterine-wall contact and self-contact, revealing largely distinct distributions across the fetal skin (median Jaccard overlap, 5.4%). Sustained episodes accounted for most contact time of both types, and self-contact involved diverse combinations of body parts. Length changes defined 32 anatomically and functionally interpretable modules among 162 modelled muscles. Using hand contact as an example, we identified muscle-length changes that varied with the contact partner and extended beyond the touching arm. By characterising movement-related conditions for tactile and proprioceptive feedback, cine-FSM provides a quantitative basis for studying multimodal sensorimotor experience during prenatal development.

---

Spontaneous whole-body movements are characteristic of early development.[1, 2] These movements generate tactile and proprioceptive feedback that may contribute to sensorimotor maturation.[3] Sensory feedback from spontaneous movements recruits developing circuits in the spinal cord and somatosensory cortex in neonatal rats.[4, 5] Such inputs participate in the interplay between spontaneous and sensory-driven activity during cortical development.[6] Altering tactile feedback during brief spontaneous movements in sleep can change spinal sensorimotor function.[7] In human neonates and infants, our previous work combining motion capture with musculoskeletal modelling revealed body-wide organisation in movement-related sensorimotor information.[8] Simulations of fetal brain–body–environment interactions have further suggested that movement-generated feedback could support cortical learning of body representations.[9] However, the physical conditions underlying this feedback remain difficult to quantify across the human fetal body in vivo.

Spontaneous movement also has clinical relevance. Movement-quality assessment can support early neurological evaluation and identification of infants at risk of cerebral palsy.[10, 11] Quantitative video analysis seeks to complement this approach.[12–14] Before birth, ultrasound has documented movements across gestation, including hand movements and self-directed contact.[15–18] Cine MRI has enabled observation of fetal motor behaviour in conjunction with brain imaging and later developmental outcomes.[19–21]

Early cine MRI studies documented fetal behaviour using one or a few thick sections, providing broad anatomical coverage but limited through-plane detail.[20] Selected in-plane kicks were subsequently combined with mechanical and musculoskeletal models to estimate limb forces.[22] Motion-corrected MRI has advanced fetal body reconstruction and anatomical analysis, whereas body models have enabled the estimation of fetal shape and pose.[23–25] Changes in muscle length provide a mechanical description relevant to proprioception.[26, 27] The remaining challenge is to quantify these changes together with skin contact while maintaining anatomical correspondence during whole-body movement.

Here we introduce cine MRI–based fetal sensorimotor mapping (cine-FSM), which combines rapid volumetric imaging with body-surface and musculoskeletal modelling. We use a low-flip-angle acquisition (Methods). Semiautomatic segmentations of approximately 1% of the acquired two-dimensional images are propagated and verified to guide body-part tracking. A statistical infant body model provides a continuous skin surface and model-defined joints, and musculoskeletal modelling estimates individual muscle lengths.[28, 29] The reconstructed surfaces can distinguish estimated uterine-wall contact from self-contact. We assessed reconstruction performance and the sensitivity of the derived measures to pose errors, characterised whole-body contact and muscle-length patterns, and examined hand contacts in relation to their partners and concurrent changes elsewhere in the body.

## 1 Results

### 1.1 Whole-body movement reconstruction

Cine-FSM reconstructed the whole-body movements of 11 third-trimester fetuses from approximately 38 min of volumetric cine MRI acquired at 2.3–3.3 volumes per second (median analysed duration, 4.2 min per fetus; range, 1.1–4.6 min; Table S1). For each fetus, tracking 14 body parts maintained anatomical correspondence. The reconstructed surfaces and model-defined joints supported estimates of uterine-wall contact, self-contact and the lengths of 162 modelled muscles on a common interpolated 24 Hz time base (Fig. 1 and Methods).

**Fig. 1.**
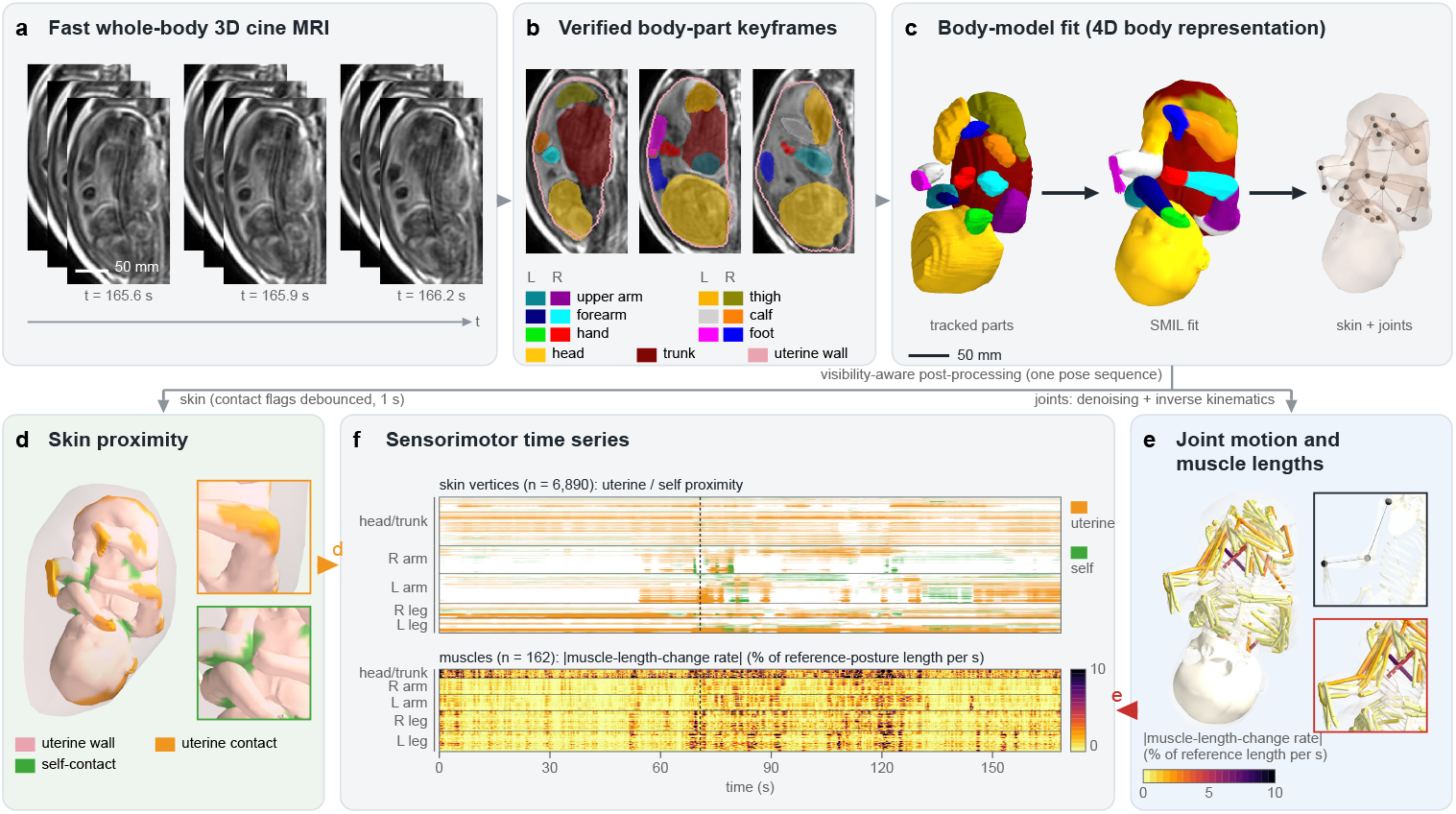
Cine MRI–based fetal sensorimotor mapping (cine-FSM). (a) Successive MRI volumes and slices within each volume. (b) Verified segmentations of the uterine wall and fetal body parts. (c) Tracked body parts, fitted SMIL surface coloured by body part, and reconstructed skin with model-defined joints. (d) Estimated uterine-wall contact (orange) and self-contact between body parts (green). (e) Joint motion and muscle lengths derived from the reconstructed pose sequence. Muscles are coloured according to the absolute muscle-length-change rate at the displayed time (100*|dL/dt|/L*_ref_ in % of reference-posture length s*^−^*^1^; 0–10, clipping higher values), which is shown as in Fig. 8b. The upper inset marks the left hip, knee and ankle joint centres of the musculoskeletal model. (f) Illustrative contact estimates and absolute muscle-length-change rates (scale of e) from fetus 7 on a shared 24 Hz numerical time base. Interpolation adds no acquired temporal information. The rates are defined as in Fig. 6.

The reconstructions captured leg extension, hand-to-face contact and trunk rotation across a range of amplitudes, with few large excursions in some recordings (Fig. 2 and Movies S1–S11). Interactive three-dimensional visualisations were generated for all 11 recordings (Data availability).

**Fig. 2.**
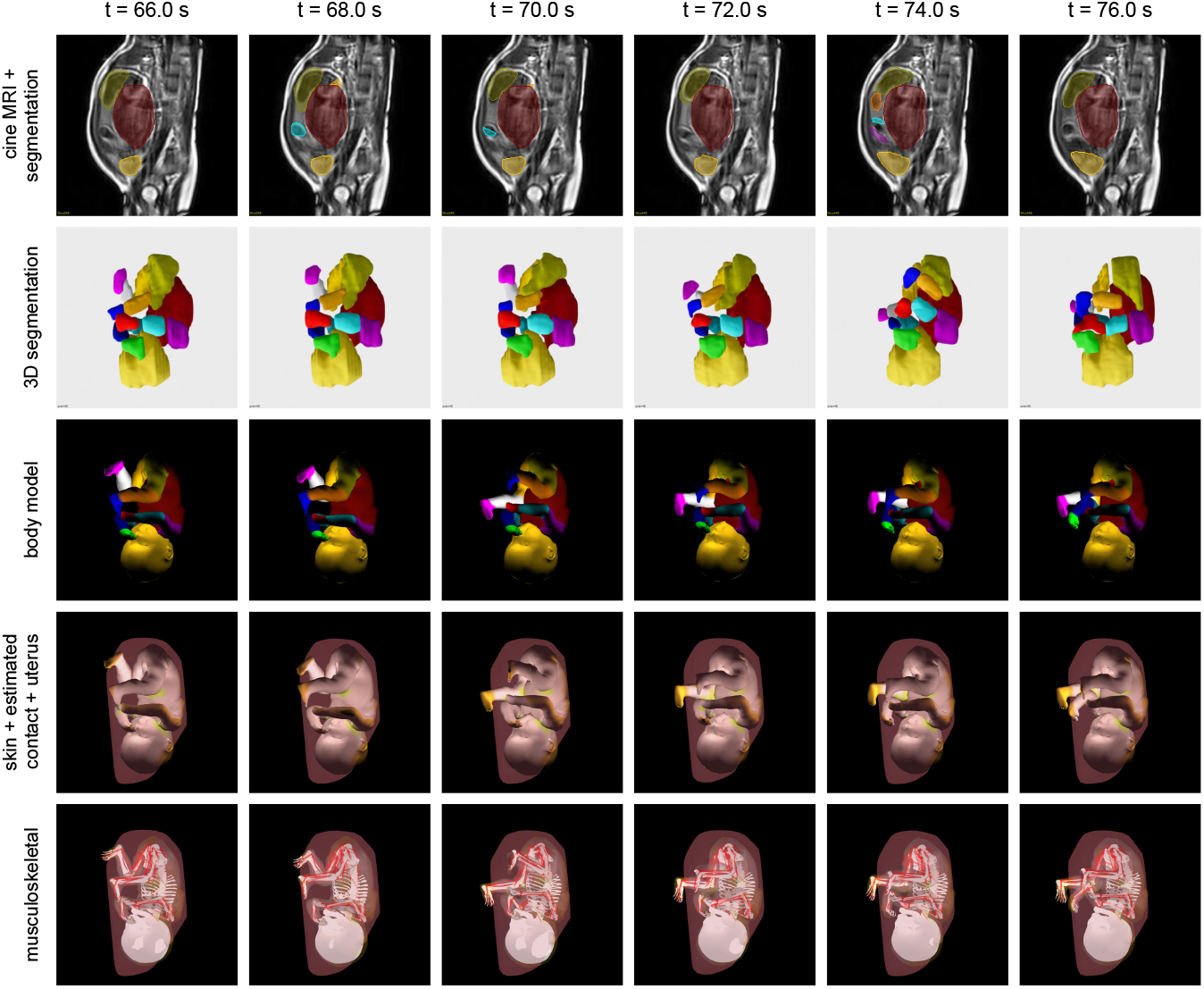
An example whole-body reconstruction. Six time points at 2-s intervals (66–76 s) from fetus 7 show movements of both legs and arms. From top to bottom, rows show cine MRI with segmentation, three-dimensional segmentation, the body model coloured by body part, skin contact flags (uterine wall, orange; self-contact, green) with the uterus, and the musculoskeletal representation. At each time point, the latter three rows use the same processed pose and camera view. Movies S1–S11 show each recording at real-time speed, and Movies S13–S15 show rotating overviews of fetuses 7, 8 and 10 with the concurrent cine MRI.

### 1.2 Reconstruction performance

Reconstructed joint configurations agreed with the manual MRI landmarks annotated with spatial uncertainty after reconstruction (22 volumes, 11 fetuses; 87 elbow or knee joint–volume pairs). The concordance correlation was 0.86, and 74% of the reconstructed elbow and knee angles lay within the annotations’ 5th–95th-percentile intervals (Fig. 3c and Fig. S2). On average, the reconstructed angles were 10.6° more flexed than the annotations, and this offset was retained in the downstream pose sequences (Fig. S2).

**Fig. 3.**
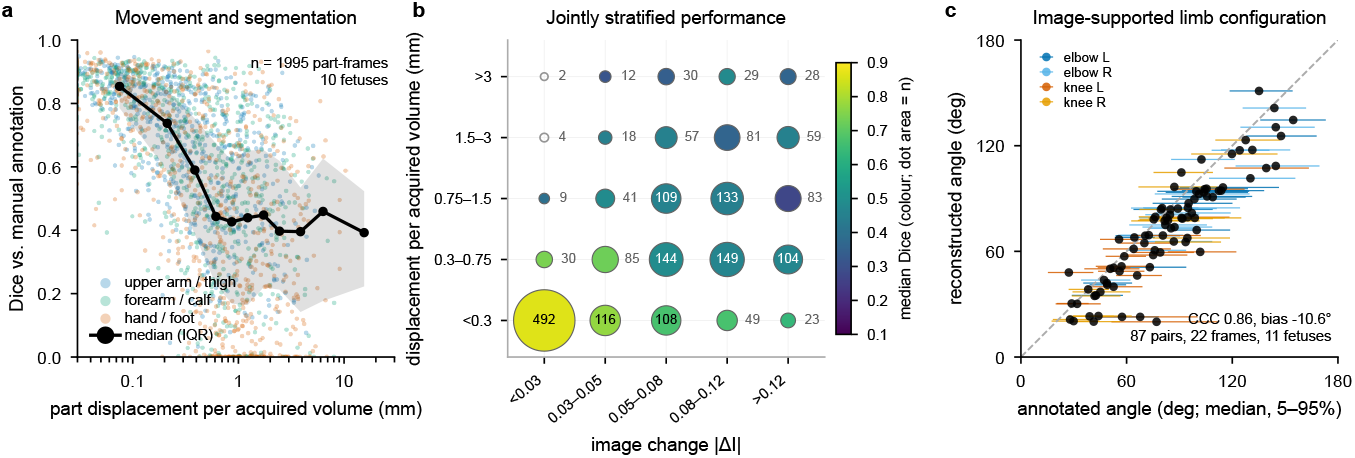
Reconstruction performance and pose-level agreement. (a) Limb Dice against manual annotation versus part-centroid displacement per frame from the nearest production keyframe (1,995 part-frames, 10 fetuses). Points represent part-frames, which are coloured by segment class. The line and band show the median and interquartile range, respectively. (b) Median Dice values stratified jointly by speed and within-part image change. Dot areas and the numbers indicate part-frame counts (open circles, fewer than 8 part-frames, no median shown). (c) Reconstructed elbow and knee configuration angles versus angles derived from manual MRI landmark annotations, with uncertainty intervals being shown (22 acquired volumes; 87 joint–volume pairs). Fig. S2 provides the full pose-level analysis, including coverage by annotation intervals before and after removal of the mean flexion bias. Panels a and b use production keyframes conditioned on manual frames, including the target. The appearance-change metric in b uses the target annotation.

The segmentation accuracy depended on the body part, movement and distance from a verified keyframe. In the production-pipeline evaluation, head and trunk Dice scores against hand-drawn masks remained comparatively stable (median, 0.82; interquartile range, 0.78–0.86; 185 cases, 10 fetuses), whereas limb Dice scores decreased with increasing distance from the nearest keyframe, from 0.75 (0.50–0.87) at 20–30 frames to 0.47 (0.26–0.71) beyond 100 frames (Fig. S1b), and declined with increasing movement speed (*ρ* = *−*0.63; Fig. 3a) and changing image appearance (Fig. 3b). Image-guided fitting between keyframes increased the limb Dice value in held-out frames from 0.68 to 0.71 (means of nine fetuses, 60 frames). Postprocessing reduced the maximum intersegment penetration by an order of magnitude, retaining 79–96% of the 90th-percentile joint speed at the acquisition rate (Fig. S1).

### 1.3 Joint kinematics and limb-movement bouts

The reconstructions provided continuous trajectories of 23 limb and lumbar joint degrees of freedom (Fig. 4b). Observed joint-angle ranges, averaged across sides, were widest at the knee and the shoulder plane of elevation and narrowest for shoulder elevation and lumbar extension, with wide variation between fetuses (Fig. 4e). Joint angular velocities exhibited bilateral and within-limb covariation. Angular velocities were positively correlated between the left and right sides for hip flexion (0.47), shoulder elevation (0.46), elbow flexion (0.40) and shoulder plane of elevation (0.25). Within-limb correlations linked shoulder rotation to forearm pronation–supination (right, 0.29; left, 0.38), right hip rotation to ankle flexion (0.35) and right hip flexion to knee flexion (0.27). Left shoulder rotation and elbow flexion were negatively correlated (*−*0.25; Spearman coefficients summarised across 11 fetuses; Fig. 4f).

**Fig. 4.**
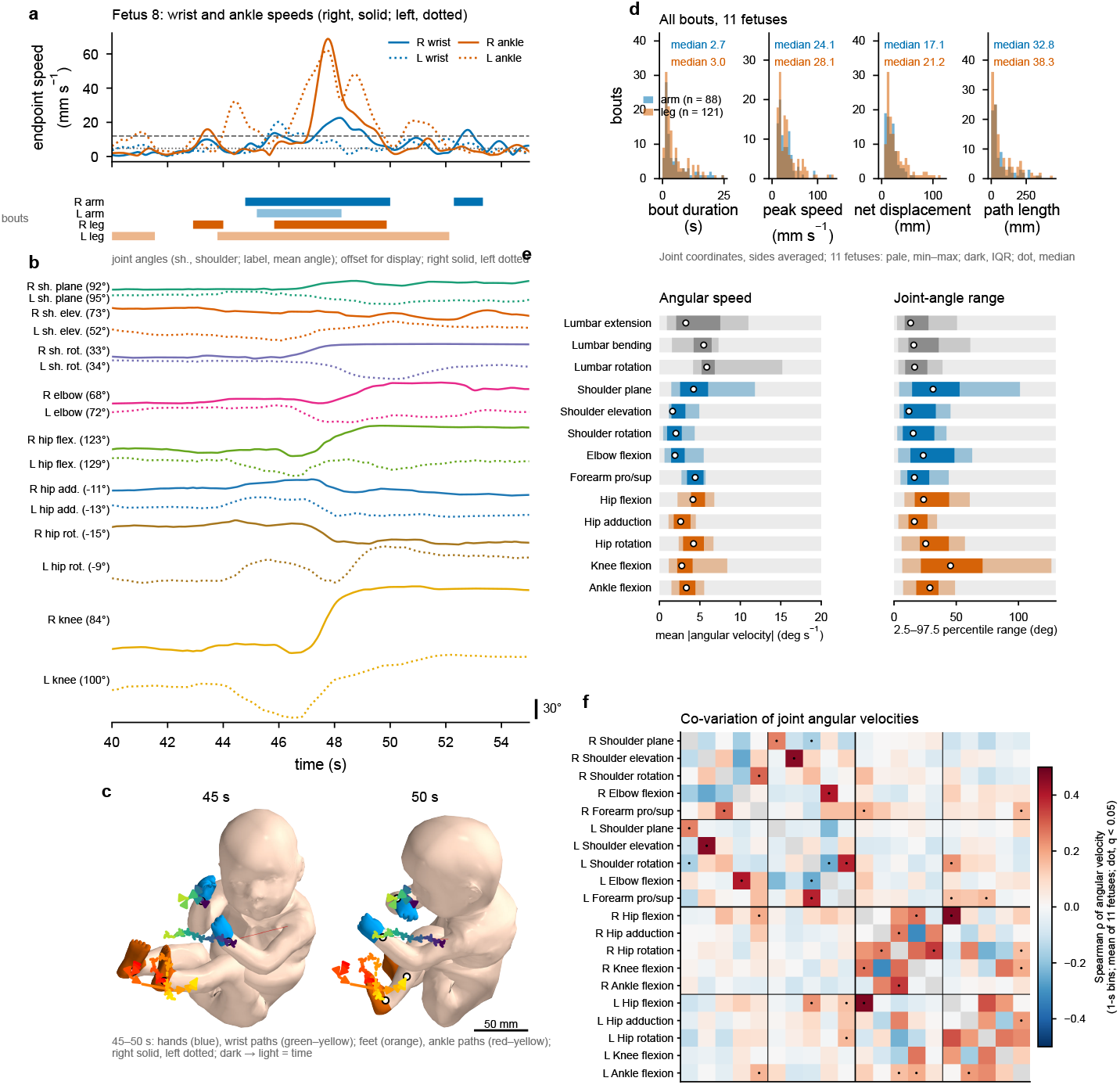
Limb-movement bouts and joint kinematics. (a) Wrist speed relative to the shoulder and ankle speeds relative to the hip in fetus 8 over 40–55 s (right, solid; left, dotted; arms, blue; legs, orange). The horizontal lines mark the 12 mm s*^−^*^1^ peak and 4.8 mm s*^−^*^1^ boundary criteria. The bars below mark accepted bouts. (b) Shoulder plane of elevation, elevation and rotation; elbow flexion; hip flexion, adduction and rotation; and knee flexion in the same interval. Traces are offset for display, with mean angles shown in the labels (scale bar, 30°). (c) Skin at 45 and 50 s, with hands coloured blue and feet coloured orange. Wrist paths progress from green to yellow, and ankle paths progress from red to yellow over 45–50 s (right, solid; left, dotted; scale bar, 50 mm). (d) Duration, peak speed, net displacement and path length of 88 arm bouts and 121 leg bouts across the 11 recordings. (e) Mean absolute angular velocity and 2.5–97.5 percentile range of 13 joint coordinates averaged across sides (pale bars, fetal minimum–maximum; dark bars, interquartile range; dots, median). (f) Spearman correlations of angular velocities in 1-s bins among ten limb coordinates on each side, which are shown as the mean values across 11 fetuses computed in Fisher z space and transformed back to the correlation scale. The black dots indicate pairs with coefficients that differed from zero across the 11 fetuses (one-sample t-test on Fisher-transformed coefficients; Benjamini–Hochberg *q <* 0.05 over 190 pairs). The limb blocks are ordered as the right arm, left arm, right leg and left leg. The criteria and global coordinate frame are described in the Methods section.

For subsequent analyses, we defined limb-movement bouts from wrist and ankle speeds relative to the shoulder and hip, respectively, using fixed speed, duration and displacement criteria (Fig. 4a and Methods). This identified 88 arm and 121 leg bouts in nine fetuses, with arm bouts in seven (Fig. S3). Arm and leg bouts had median durations of 2.7 and 3.0 s, respectively; median peak speeds of 24 and 28 mm s*^−^*^1^; and median net displacements of 17 mm (interquartile range, 11–25 mm) and 21 mm (13–38 mm). Their path lengths exceeded the net displacement (medians, 33 and 38 mm; maxima, 350 and 430 mm; Fig. 4d).

### 1.4 Whole-body contact maps

We estimated contact from the geometric proximity of reconstructed surfaces, distinguishing uterine-wall contact from self-contact (Methods). The time-averaged proportion of skin vertices in uterine-wall contact ranged from 15% to 43% across the fetuses (mean, 26%), compared with 3–17% for self-contact (mean, 9%; Fig. 5a–c). The contact types occupied largely distinct skin regions (median Jaccard overlap, 5.4% across the fetuses), with spatial separation persisting across occupancy thresholds of 5–20%. The relative contact extent remained similar when the proximity threshold varied from 2 to 16 mm, whereas individual contact-onset detections were threshold-dependent (Fig. S4).

**Fig. 5.**
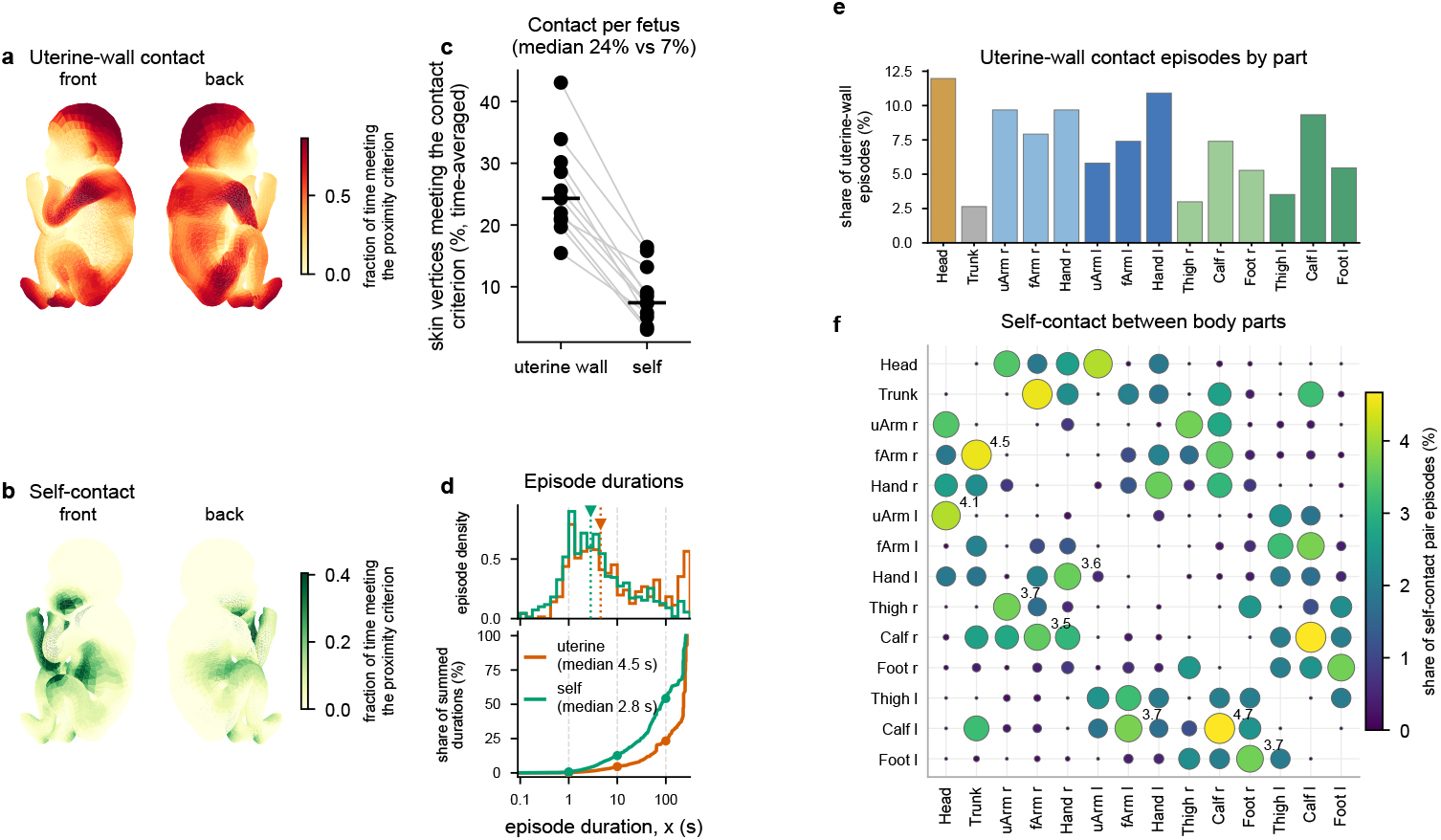
Spatial and temporal distributions of estimated skin contact. (a, b) Uterine-wall contact and self-contact occupancy on the skin (averaged across 11 fetuses) in front and back views. Occupancy is the fraction of time meeting the geometric proximity criterion. (c) Each fetus’s time-averaged fraction of skin in proximity. (d) Normalised episode-density histogram (upper) and cumulative proportion of summed episode durations from episodes shorter than *x* (lower) pooled across body parts and fetuses on a logarithmic duration axis. The median episode durations were 4.5 s for uterine-wall contact and 2.8 s for self-contact. (e) Proportions of uterine-wall contact episodes by body part (each part’s episodes divided by the 568 part-level episodes pooled across fetuses). (f) Proportions of self-contact episodes for body-part pairs (each pair’s episodes divided by the 1,499 pair episodes pooled across fetuses), which are shown as a symmetric dot matrix in which the dot area and colour both encode the proportion, with each undirected pair being displayed in both orientations but counted once. The eight largest proportions are labelled. For d and e, a body part was in contact when at least 5% of its vertices met the criterion, with gaps shorter than 0.25 s being merged. Panel f instead uses retained part-pair episodes after 0.5 s of debouncing and the duty-cycle/repetition criteria in the Methods section. These body part and pair summaries use different denominators and differ from individual-vertex dwell times, merged hand-contact events and pooled hand-frame occupancy (Results and Methods). Estimates use temporally processed skin and filtered flags (Methods) and do not measure contact force or skin deformation.

Long episodes accounted for most of the summed contact duration. At the bodypart level, the pooled median durations were 4.5 s for uterine-wall contact and 2.8 s for self-contact, but episodes shorter than 10 s contributed only 5% and 13% of the respective summed durations, and those shorter than 100 s contributed 23% and 54% (Fig. 5d). Among 568 uterine-wall episodes pooled across body parts and fetuses, the largest proportions involved the head (12.0%), left hand (10.9%), and right upper arm and right hand (9.7% each; Fig. 5e). Recurrent self-contact included calf–calf (4.7% of 1,499 pair episodes), right forearm–trunk (4.5%), head–left upper arm (4.1%) and left forearm–left calf (3.7%; Fig. 5f).

The hands, a focus of fetal behavioural research,[16–18] were in contact for much of the time. Across the sampled frames from both hands of the 11 fetuses, self-contact with the head or arm segments, including the opposite hand, occurred in 41% of the hand frames, and uterine-wall contact occurred in 44% of the frames (median fractions across fetuses of 49% and 33%, respectively). Episodes lasting at least 10 s accounted for 85% and 87% of the respective contact-positive hand frames (Methods).

### 1.5 Whole-body muscle-length patterns

We estimated length trajectories for 162 modelled muscles from the reconstructed joints, normalising each muscle’s length to its value in a common reference posture (Fig. 6a and Methods). During these limb-movement bouts, the mean positive lengthening rate in muscles of the moving limbs was 1.06% of reference length per second (median across the nine fetuses with detected bouts; Fig. 6c and Fig. S7); over the full recordings, muscle lengths varied by a median of 8.1% of reference length (1.7–18.7% across fetuses; Fig. S5).

**Fig. 6.**
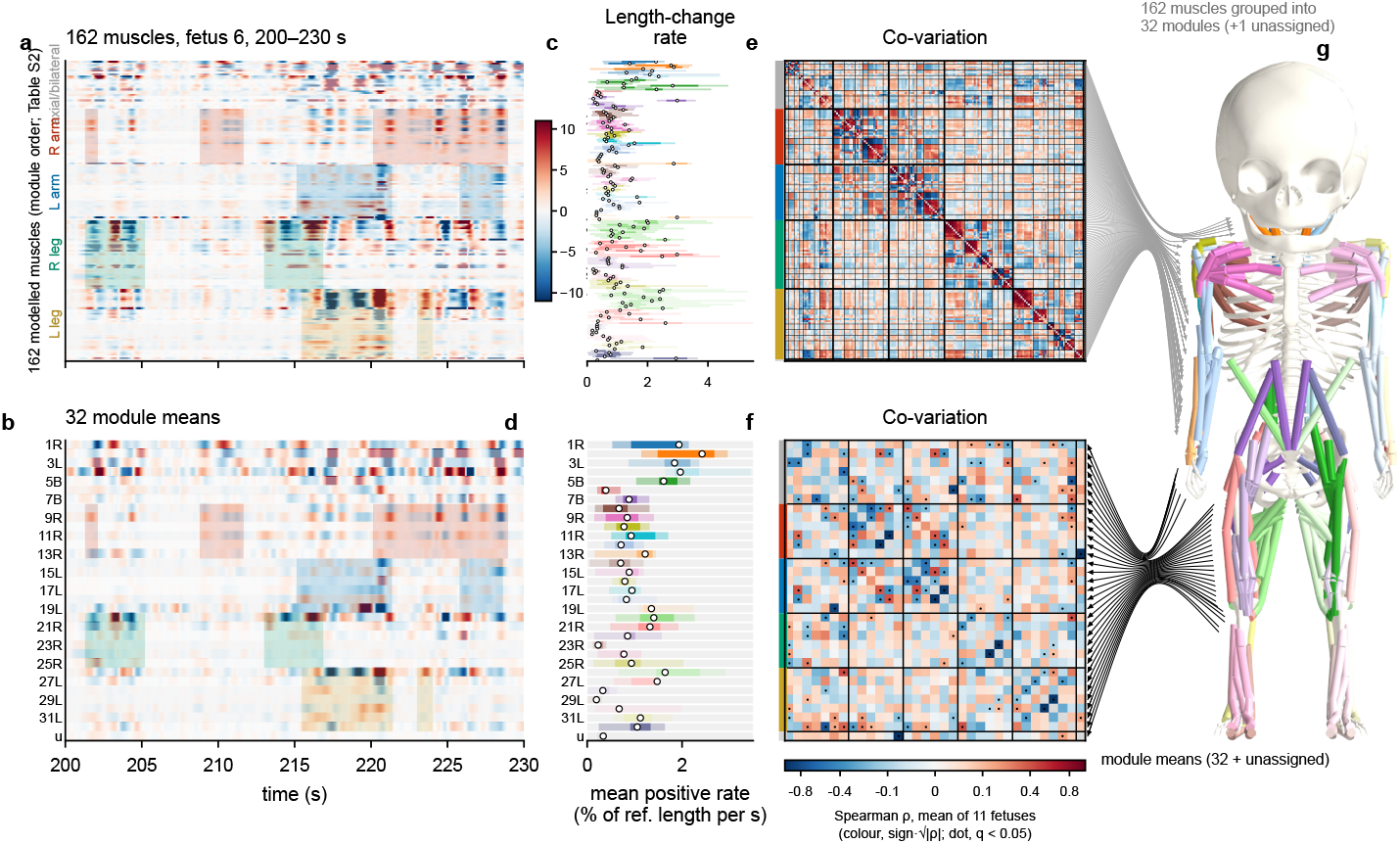
Muscle-length changes and data-driven modules. (a) Signed length-change rate of all 162 muscles in a 30-s interval of fetus 6 (200–230 s; % of the reference-posture length s*^−^*^1^; red, lengthening; blue, shortening) ordered by the 32 data-driven modules (Table S2), spanning from axial or bilateral groups to the four limbs, with the unassigned muscle being shown last. Colour saturates at *±*11.0% s*^−^*^1^, which is the 98th percentile of the absolute rate in the interval. Shaded rectangles mark the limb-movement bouts over the rows of that specific limb’s muscles (in the colours of Fig. 7a). Neck and trunk rows are unshaded because bouts are defined from limb endpoints (Methods). (b) The same rates as the 32 module means plus the unassigned muscle (33 rows; R, L and B denote right, left and bilateral membership; every second row labelled), with the scale and shading of a being used. (c) Mean positive lengthening rate within bouts for each muscle (in the same order of a) for the nine fetuses with detected bouts (Fig. S3; Methods). Pale bars, fetal minimum–maximum; dark bars, interquartile range; dots, median. Nonlengthening samples contribute a value of zero. The colours denote the module. The left modules share the hue of their right counterpart in a lighter tint, and the unassigned muscle is coloured grey. The 14 modules with mean stability scores of less than 0.8 are shown in pale colours; these modules are assigned. Panel c shows the distribution across fetuses for each muscle. For the main-text summary, eligible limb-muscle samples were pooled within each fetus before averaging, and the median was then obtained across the nine fetuses. (d) Positive rate by module within bouts (with conventions as shown in c). (e) Muscle-level Spearman correlations of signed rates in 1-s bins, which are shown as the means across 11 fetuses computed in Fisher z space and transformed back to the correlation scale, with the limb colour of each row on the left. (f) The same approach was used for module-mean rates, which were computed within fetuses (32 modules and the unassigned muscle, diagonal hidden). The black dots indicate module pairs with coefficients that differed from zero across the 11 fetuses (one-sample t-test on Fisher-transformed coefficients; Benjamini–Hochberg *q <* 0.05 over the 528 pairs). The correlation colours are proportional to sign(*ρ*) 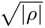, with ticks in *ρ*. The black lines separate the axial/bilateral and four limb blocks. (g) Module membership on the musculoskeletal model, as drawn in an upright posture with all joint coordinates at zero for display. The grey lines lead from the 162 rows of e towards the model and merge into one arrow per module (32 modules and the unassigned muscle); the black arrows lead from the model to the 33 rows of f, which summarises the module means. Panels c and d use bout frames of nine fetuses, whereas e and f use the full recordings of all 11 fetuses, as did the module definition. Neck and trunk configurations are less constrained by the images than limb configurations. Shared joint geometry can produce these patterns without muscle activation or neural coordination. Because membership was derived from the same recordings, within-module covariation is descriptive.

To determine whether the 162 muscle-length trajectories contained shared patterns, we grouped the muscles by correlations in their length-change rates without using anatomical labels. This yielded 32 modules of 2–11 muscles and one unassigned muscle (Table S2 and Methods). Within a module, muscle rates were strongly correlated (median, 0.65), whereas correlations between modules were weak (median absolute value, 0.076). The mean signed rate of the other muscles in that module accounted for a median of 58% of an individual muscle’s rate variance (47–71% across fetuses). Therefore, the modules provided a compact description of shared musclelength changes across the body. The mean resampling stability score was 0.81 across the 161 assigned muscles, with 98 muscles scoring at least 0.8. Sensitivity to analysis settings is reported in Table S3.

The modules aligned with anatomical and functional groupings. Elbow flexors and extensors formed separate modules on each side, as did deep plantar flexors and dorsiflexors, with opposing length changes (*ρ* = *−*0.97 and *−*0.95 at the elbow, and *−*0.98 and *−*0.95 at the ankle; Fig. 6f). The neck, trunk and hip groups also had interpretable functional labels, although some groups were mixed (Table S2). No module combined arm and leg muscles, and 28 of 32 modules were confined to one side. Each of the six right-arm and six right-leg modules had a largely matching left counterpart (mean mirrored-membership Jaccard overlap of 0.93 and 0.90, respectively), and the within-limb correlation structures were similar across sides (*r* = 0.90 and 0.89, respectively). The whole-recording summaries in Fig. 7d,e use predefined anatomical regions, and the bout-centred analysis in Fig. 7f uses muscle modules.

**Fig. 7.**
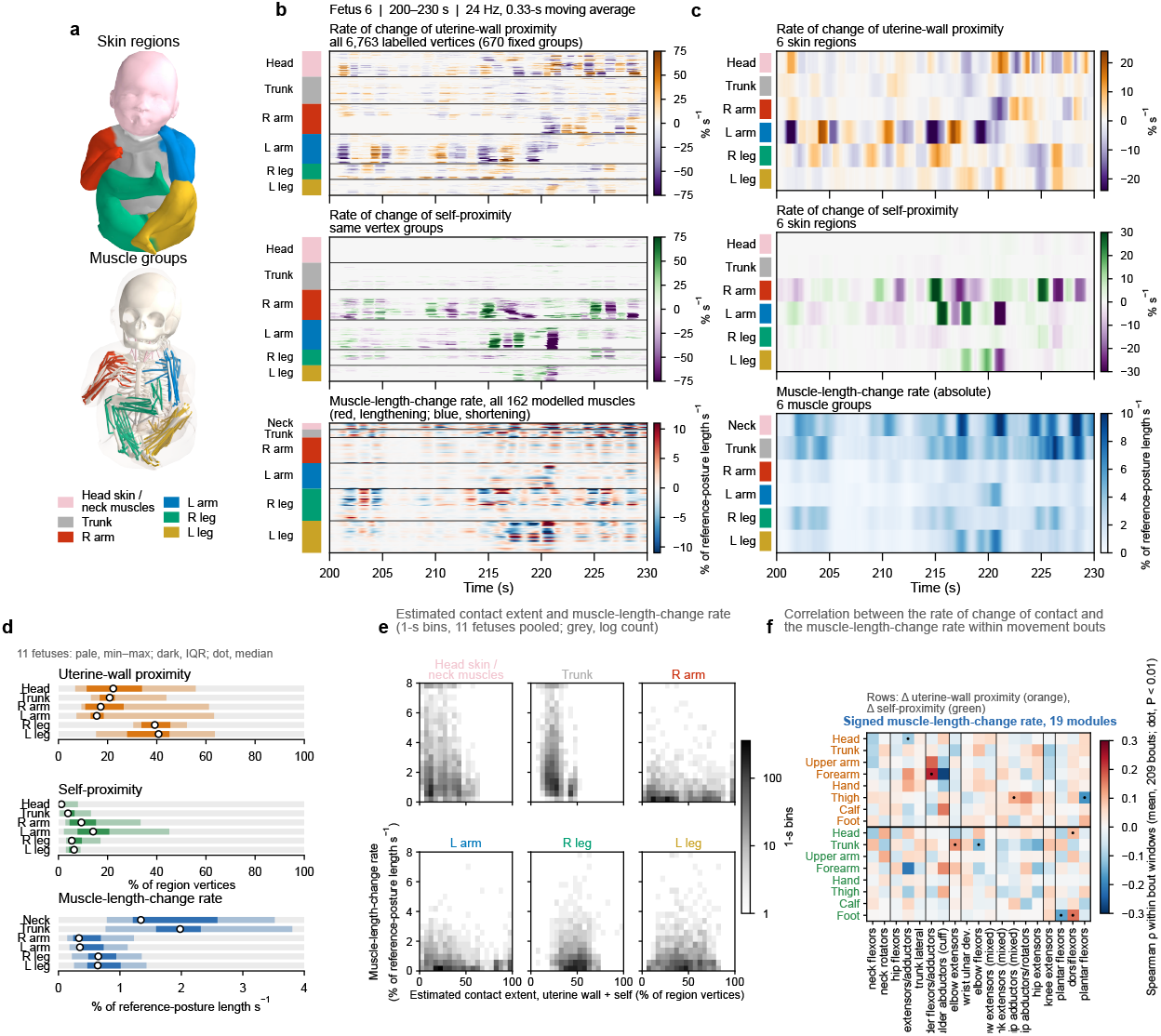
Contact estimates and muscle-length changes across scales. (a) Six skin regions and six muscle groups, as shown on fetus 10. Panels b and c show fetus 6. Matching colours link each skin region to its anatomically corresponding muscle group, with the head skin paired with the neck muscles. The regional display order is head/neck, trunk, right arm, left arm, right leg and left leg. (b) The 200–230 s interval of fetus 6 (also shown in Fig. 6a,b). The rate of change in contact is shown for 670 fixed region-ordered groups of the 6,763 labelled skin vertices, alongside signed muscle-length-change rates for all 162 muscles. The contact fractions were differentiated on the 24 Hz numerical time base after a 0.33 s moving average (orange–purple, uterine-wall contact; green–purple, self-contact). The muscle scale is clipped at *±*12% of the reference length s*^−^*^1^ (0.8% of the displayed cells). (c) The same interval summarised by skin region as the rate of change in contact and by muscle group as the mean absolute muscle-length-change rate. (d) Regional recording means across 11 fetuses (pale, minimum–maximum; dark, interquartile range; dot, median). (e) Pooled 20 *×* 20 joint histograms of estimated contact extent and the corresponding muscle-length-change rate using 2,252 valid 1-s bins from 11 fetuses. Greyscale represents log counts. The extent is the sum of the wall and self-fractions (clipped at 100%) and approximates their union. Pooling does not isolate within-fetus association. (f) Spearman correlations within 10 s windows centred on each of the 209 limb-movement bouts in nine fetuses. Rows represent the rate of change in contact on eight body parts on the moving side, with uterine-wall contact being situated above and self-contact being situated below. Columns represent the signed muscle-length-change rates of the 19 bilaterally folded modules on that side (grouped by region). Displayed values are means across bouts computed in the Fisher z space and transformed back to the correlation scale. Cell-wise tests included eight fetuses in 298 cells, seven fetuses in four cells and six fetuses in two cells (Source Data). Dots mark cells with nominal *P <* 0.01 at the fetus level (nine of 304; none survived Benjamini–Hochberg correction). The whole-recording partial-correlation matrix among the 18 regional series is reported in Fig. S8 and describes pair-specific conditional associations. All muscle-length-change rates are normalised to the reference length. Regional muscle summaries average the absolute rates of individual muscles, whereas the fine-scale trajectories and bout-centred module analysis use signed rates. The contact fractions use labelled regional vertices as denominators. The geometric criterion indicates estimated contact (not confirmed physical touch).

### 1.6 Concurrent contact and muscle-length changes

We examined how contact and muscle-length changes were related across the body using six anatomically corresponding skin regions and muscle groups (Fig. 7a–c).

Regional contact levels differed between the arms and legs. Uterine-wall contact was more extensive in the legs (median fractions, 39–41% versus 16–17% in the arms), whereas self-contact was more extensive in the arms (9–14% versus 5–6% in the legs; Fig. 7d). In pooled regional distributions, similar contact extents were accompanied by a wide range of muscle-length-change rates (Fig. 7e).

Contact series represented rates of change in uterine-wall and self-contact fractions, whereas regional muscle rates averaged absolute length-change rates. In partial correlations adjusted for muscle-length-change rates in other regions, temporal associations were more evident among contact measures and among muscle groups than between contact and muscle measures (Methods and Fig. S8). Muscle-length-change rates were positively associated between the arms and between the legs (median partial correlations, 0.45 and 0.42; positive in ten and 11 fetuses, respectively), and the two contact types tended to change in opposite directions within regions (median, *−*0.17; negative in all 11 fetuses). None of the 72 skin–muscle pairs reached *P <* 0.01 across fetuses. All 17 associations surviving correction over 153 pairs were skin–skin or muscle–muscle (Benjamini–Hochberg *q <* 0.05; Fig. S8).

To examine contact–muscle associations around individual movements, we analysed body-part contact changes and signed muscle-module rates in 10 s windows centred on 209 limb-movement bouts in nine fetuses (Fig. 7f). Nine of the 304 part–module cells reached nominal *P <* 0.01 in the fetus-level analysis, but none survived multiple-testing correction (Methods). With the two contact types merged, two of 152 cells met the nominal threshold and none survived correction (Fig. S9).

### 1.7 Hand contact within its whole-body context

We focused on hand-contact events to examine how local contacts with different partners were embedded in ongoing whole-body movement. We identified 102 events in nine fetuses, including 41 uterine-wall contacts and 61 self-contacts with the head (21) or an arm (40: the opposite hand, 13; the opposite forearm, 19; and the same-side upper arm, 8), and separately analysed these three contact families (Methods).

A left-hand-to-head event in fetus 10 illustrates concurrent contact and muscle-length changes across the body (onset, 184.3 s; Fig. 8a–c and Movie S12). The minimum skin gap decreased from approximately 15 mm at *−*1.5 s to 3.5 mm at +0.5 s, whereas the fraction of hand vertices within 10 mm of the head increased after onset. Regional magnitudes average absolute module-mean signed rates (Methods), and module names follow predominant actions (Table S2). Pooled across 102 events, magnitudes were greater in all regions before contact (*−*1.5 to *−*0.4 s) than in nearby control windows in the same recording (differences, +0.29 to +1.18% of the reference length s*^−^*^1^; all *q* = 0.001). Neck-flexor signed rates were higher, and neck-rotator rates were lower (*q ≤* 0.015), whereas signed-rate differences in the seven arm modules were small and nonsignificant (absolute differences *≤* 0.11% of the reference length s*^−^*^1^; *q ≥* 0.14).

**Fig. 8.**
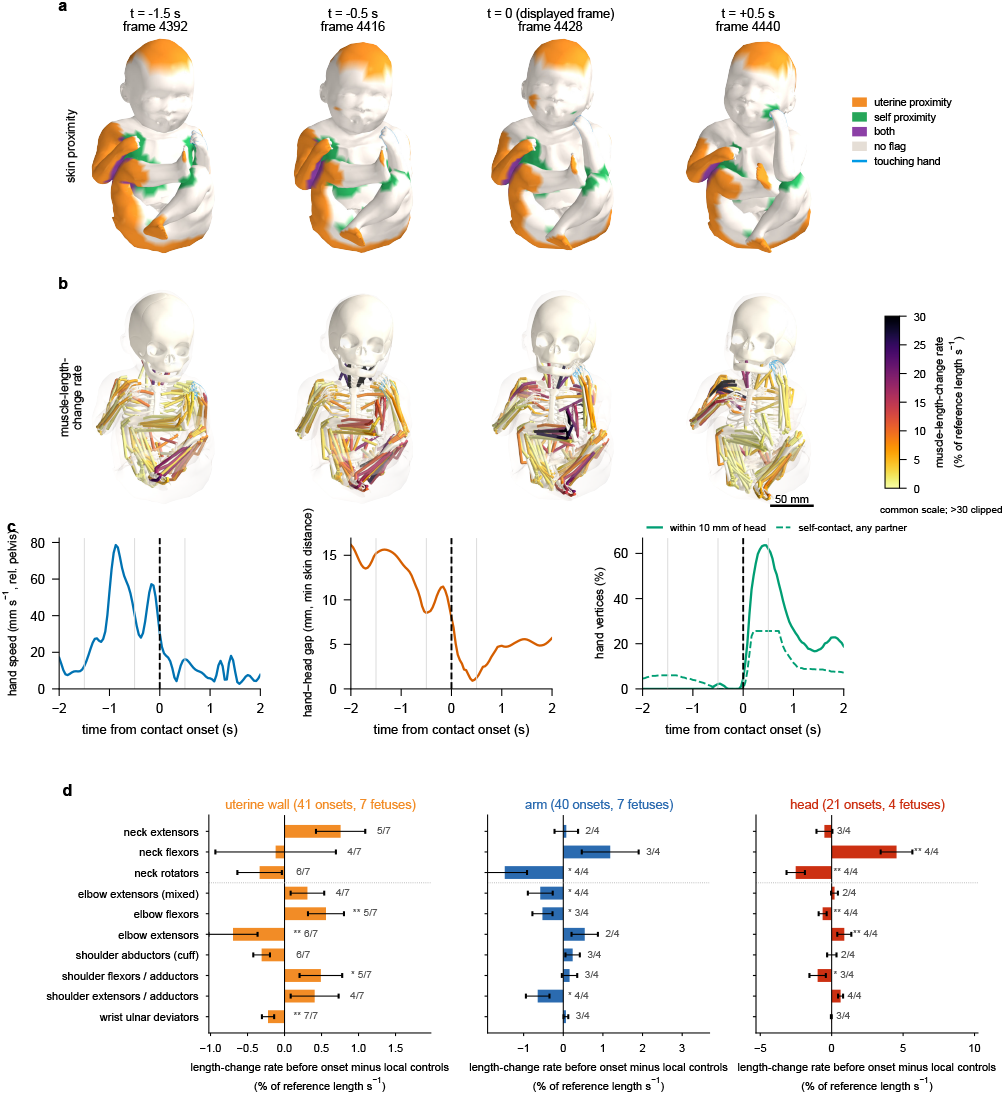
Hand contact in its whole-body context. (a–c) An illustrative hand-to-head contact. Fetus 10, left hand (onset, 184.3 s on the 24 Hz numerical time base). The displayed *t* = 0 frame (4428) is 0.17 s later than the listed onset frame; this offset is shorter than one acquired-volume interval. The event was selected for frontal visibility. (a) Skin at *−*1.5, *−*0.5, 0 and +0.5 s, as coloured by proximity flags (uterine wall, orange; self, green; both, purple; no flag, cream). Self-proximity does not identify the partner. The hand has a cyan outline. The camera, projection and scale are shared (bar, 50 mm). (b) The same time points with the 162 muscles coloured by absolute length-change rate (100*|dL/dt|/L*_ref_ , in % of the reference length s*^−^*^1^) using a shared 0–30 scale and clipping higher values. Bones provide an anatomical reference. Faint skin indicates orientation. Auxiliary translations were locked at zero. The model has no intrinsic hand or facial muscles. (c) Hand speed relative to the pelvis, minimum hand–head skin gap, and the fraction of hand vertices within 10 mm of the head (solid) or carrying a self-contact flag with any partner (dashed) over a range of *−*2 to +2 s. The 10 mm display criterion differs from the onset threshold. Onset uses the fifth-smallest gap with hysteresis (rather than the minimum gap plotted here). The dashed black line indicates onset, and grey lines indicate the displayed times. Movie S12 shows the same event from *−*2 to +2 s at one-quarter speed, with the skin and proximity flags (as shown in a) situated beside the muscles on the bones of the scaled model (as shown in b). (d) Differences in preonset signed module-mean muscle-length-change rates of the touching arm and neck relative to local control windows, as shown by contact family (uterine wall, 41 onsets in seven fetuses; arm, 40 onsets in seven fetuses; head, 21 onsets in four fetuses). For each onset, the rate was averaged over a range from *−*1.5 to *−*0.4 s, and the mean value of up to four local control windows 6–14 s away was subtracted. Bars show the event-weighted mean onset–control difference; error bars show the s.e.m. of the fetus-specific mean differences. Positive differences indicate higher signed rates before onset than in local control windows (not necessarily indicating absolute lengthening). Asterisks mark Benjamini–Hochberg *q <* 0.05 (two) or *q <* 0.10 (one) from the permutation of the onset label within each onset’s set of windows (10,000 permutations), corrected over six regional rate magnitudes, one whole-body summary and ten signed module rates within each contact family. Fractions report direction agreement among fetuses with at least two onsets as the number agreeing divided by the number eligible.

When contacts were separated by partner, the elbow-associated differences from local control windows had opposite signs for uterine-wall and head contact (Fig. 8d). Before uterine-wall contact, the signed rates were higher than in local control windows for elbow flexors and lower for elbow extensors and wrist ulnar deviators (differences, +0.56%, *−*0.70% and *−*0.22% of the reference length s*^−^*^1^; *q* = 0.013, 0.041 and 0.002, respectively; same direction in five to seven of seven fetuses).

Before head contact, the elbow-associated differences had the opposite signs, with lower flexor and higher extensor rates (*−*0.63% and +0.88% of the reference length s*^−^*^1^; *q* = 0.012 and 0.009, respectively). Neck-flexor rates were higher, and neck-rotator rates were lower, than in local control windows (same direction in all four fetuses; *q ≤* 0.012). Regional rate magnitudes also exceeded local control values in the trunk and the whole-body summary (+3.19% and +1.15% of the reference length s*^−^*^1^, respectively; both *q* = 0.001).

Before arm contact, signed rates were lower for elbow flexors, shoulder extensors and adductors, and neck rotators than in local control windows, without meeting the within-family *q <* 0.05 criterion (*q* = 0.091; same direction in three to four of four fetuses with at least two onsets).

The onset–control increase in the trunk rate magnitude was greater before head contact than before uterine-wall contact (between-family difference, 3.2% of the reference length s*^−^*^1^; *q* = 0.017; same direction in all four fetuses contributing to both families). The corresponding head–arm difference was 3.1% of the reference length s*^−^*^1^ (*q* = 0.08). The elbow-related differences between uterine-wall contact and head contact reached only nominal significance (*P* = 0.02; *q* = 0.10; Table S4).

Hand contact occurred amid continuing activity elsewhere in the body. During the first second of self-contact, a median of ten other body-part pairs were in self-contact and 11 of 14 body parts met the uterine-wall criterion. Nearby control times had comparable contact counts. In the preceding 2 s, the touching arm contributed 16% of the summed absolute normalised muscle-length change (Fig. S6).

### 1.8 Robustness of the sensorimotor maps

To assess sensitivity to pose errors, we perturbed limb-joint angles in two fetuses using the landmark-derived flexion offset and residual spread, then recomputed contact and muscle-length estimates (Methods and Fig. S10). Under a constant offset, muscle-length-change rates remained highly correlated with the unperturbed rates (correlation, 1.00), whereas time-averaged contact fractions shifted by 1–4 percentage points. With slowly varying errors (s.d. of 8.5°, below 0.1 Hz), regional and module length-change rates remained highly correlated with the unperturbed rates (correlations, 0.91–1.00 and 0.95, respectively), although their magnitudes increased by 11–13%. Uterine-wall contact time courses were less affected than self-contact time courses (correlations with the unperturbed series, 0.72–0.85 versus 0.53–0.78, respectively). In the faster-error scenario (joint-angle errors band-limited below 1 Hz), length-change rates increased to 2.5 times the unperturbed values.

Under slowly varying errors, correlations between the perturbed and unperturbed bout-centred contact–muscle matrices were 0.79 and 0.76 for the two fetuses, compared with 0.13 between their unperturbed matrices. With the onset and local control windows held fixed, the preonset responses of the same 38 hand-contact events remained highly correlated with the unperturbed responses (correlation, 0.99), with sign agreement in 93–98% of the event–measure pairs. In the faster-error scenario, the correlations were 0.94–0.96, and the sign agreement was 87–88% (Fig. S10).

Each within-family effect in Table S4b retained its direction in every leave-one-fetus-out run. Depending on the measure, *q* remained below 0.10 in 6–7 of 7 runs for uterine-wall contact, 3–4 of 4 for head contact and 2–4 of 7 for arm contact.

## 2 Discussion

Previous studies have characterised fetal movement and the biomechanics of selected movements,[1, 15–17, 20–22] whereas recent body models have enabled three-dimensional shape and pose tracking.[25] Cine-FSM combines volumetric cine MRI with body-surface and musculoskeletal modelling to reconstruct whole-body movement in three dimensions over time and to estimate skin contact and length changes in 162 modelled muscles. This approach preserves the spatial and temporal relationships among movement, skin contact and muscle length across the fetal body.

Joint reconstruction captured both the extent of individual joint movements and their covariation across limbs (Fig. 4e,f). Extracting bouts from continuous trajectories further allowed contact and muscle-length changes to be examined around individual movements (Fig. 4a–d).

Persistent contact and local contact transitions coexisted across the body. Although many episodes were brief, episodes occurring for at least 10 s accounted for most of the summed episode durations. Local onsets occurred while contact continued elsewhere, so counts of isolated events cannot capture the concurrent contact pattern. Self-contact and uterine-wall contact occupied largely distinct skin regions, with diverse bodypart pairs involved in self-contact. Postnatal studies link early tactile experience to somatosensory processing[30] and show that tactile training can facilitate reaching towards touched body locations.[31] By mapping both contact types across space and time, cine-FSM provided the whole-body context needed to interpret individual contacts.

Musculoskeletal modelling enabled estimates of muscle lengths and their changes, providing a mechanical description relevant to fetal proprioception.[26, 27] Correlation-based grouping organised complex whole-body muscle-length changes into anatomically and functionally interpretable modules. Shared joint geometry and covarying motions can generate such structure.[32] Our findings complement sensory-synergy analyses based on estimated muscle-length changes[33] and postnatal research on body-wide sensorimotor organisation.[8] Within-limb covariation and opposing changes, together with similar module organisation on both sides, suggest structured mechanical conditions for proprioceptive feedback during fetal movement (Fig. 6).

Hand contact illustrates the importance of the contact partner. Pooled regional rate magnitudes increased before contact relative to nearby control windows in the same recording, whereas signed-rate differences in arm modules were small. Separating contacts by partner identified elbow-related differences consistent with relative shifts towards extension before uterine-wall contact and flexion before head contact. Direct comparisons supported a greater onset–control increase in trunk rate magnitude before head contact than before uterine-wall contact (Fig. 8 and Table S4). Evidence for arm contact was weaker. These findings extend ultrasound descriptions of differentiated hand movements and hand–mouth coordination[16, 17] to accompanying muscle-length changes and contacts elsewhere (Fig. S6).

Advances in behavioural measurement and biomechanical modelling are enabling new insights into sensorimotor control by combining experiments with simulation. A virtual-rodent control network trained to reproduce behaviour predicted aspects of neural activity,[34] while NeuroMechFly v2 used multisensory feedback to investigate closed-loop control.[35] Cine-FSM could supply subject-specific movement, contact and muscle-length estimates for comparison with our earlier simulations coupling brain, body and uterine environment.[9] Comparing simulated and reconstructed patterns could guide model refinement and generate predictions for subsequent measurements. This iterative exchange could support model-based tests of hypotheses about prenatal neural development and body-representation learning. A common musculoskeletal representation could also support prenatal–postnatal comparisons of patterns that persist or change as the body and environment develop.[8]

Pose-error sensitivity highlighted the importance of temporal fidelity alongside spatial accuracy. In two fetuses, slowly varying joint-angle errors (band-limited below 0.1 Hz) largely preserved muscle-length-change patterns, bout-centred associations and precontact muscle-rate differences evaluated in fixed onset and control windows, whereas faster errors inflated rate magnitudes (Fig. S10). Relative contact extent across fetuses also remained similar across proximity thresholds of 2–16 mm (median Spearman *ρ* across thresholds, 0.92 for self-contact and 0.98 for uterinewall contact), although onset detections were threshold-dependent (Fig. S4). Distal segmentation errors and systematic flexion bias limit anatomical accuracy. Implicit neural representations and scanner-integrated workflows have advanced structural fetal reconstruction.[36, 37] Adapting these structural methods to dynamic imaging requires preserving movement trajectories and event timing within the acquired spatial and temporal resolution. Improvements in imaging resolution, image contrast and articulated body models could eventually extend this approach to finer movements, including those of the fingers and eyes, and to the sensory conditions they create.

Shared model geometry and reconstruction errors may contribute to within- and cross-modal associations. Bout-centred associations remain exploratory, with none surviving multiple-testing correction in the fetus-level analysis (Fig. 7f). Muscle-length trajectories and surface proximity describe mechanical conditions rather than muscle-spindle or cutaneous afferent firing.[26, 27] Evaluating their physiological relevance requires complementary neural measurements. The observed patterns do not establish common neural drive or anticipatory control. Fetal fMRI has been used to demonstrate associations between eye movements and brain networks,[38] detect auditory blood-oxygen-level-dependent (BOLD) responses[39] and link scan-level head movements to network dynamics.[40] Integration with time-aligned cine-FSM could extend studies of prenatal sensorimotor maturation while requiring BOLD-sensitive acquisition, separation of movement-related signals from motion artefacts and consideration of fetal haemodynamics.

Generalisability is limited by short third-trimester recordings from 11 clinically referred pregnancies, with unequal event counts and four to seven fetuses per contact family. Assessing prognostic value requires longitudinal follow-up with postnatal motor and cognitive assessments. Associations of fetal functional connectivity with later motor ability[41] and of cine MRI movement quality and brain findings with developmental outcomes[21] motivate testing whether cine-FSM adds information beyond structural imaging and conventional movement assessment. Such evaluations should account for gestational age, clinical conditions, activity and posture.

Cine-FSM reconstructs and visualises fetal whole-body movement in three dimensions while quantifying the accompanying skin contact and muscle-length changes. By integrating these measures across the body on a common time base, this approach extends the analysis of individual movements and contacts to the whole-body physical patterns underlying prenatal sensorimotor experience.

## 3 Methods

### 3.1 Study participants and image acquisition

#### Ethics and participants

Eleven singleton pregnancies referred to Kyushu University Hospital for clinically indicated structural MRI were included. The cine acquisition was added to the clinical examination. Gestational age at imaging ranged from 29 weeks 6 days to 35 weeks 1 day (median, 33 weeks). The imaging indication was fetal in five pregnancies (congenital diaphragmatic hernia in three, a fetal thoracic mass and a fetal pelvic mass) and maternal or placental in six (uterine fibroids in three, complete placenta praevia, suspected placenta accreta and suspected vasa praevia). The study was approved by the Institutional Review Board of Kyushu University (approval no. 24014-00, 18 April 2024), with The University of Tokyo participating as a collaborating institution under the same central review, and written informed consent was obtained from each participant. Participant characteristics and acquisition parameters are listed in Table S1.

#### Cine MRI acquisition

We used a low-flip-angle three-dimensional balanced steady-state free precession acquisition (bSSFP, balanced fast field echo) developed by the study team. Lowering the flip angle reduces radiofrequency power deposition and allows shorter repetition and echo times within a whole-body averaged specific absorption rate limit of 2 W/kg.

Recordings used a 1.5 T scanner (Achieva dStream; Philips Healthcare) with anterior and posterior coils or a Flex-L coil with a posterior coil.

For all study acquisitions, the flip angle was 25° and the SENSE acceleration factor was 3 *×* 2 (phase *×* slice). TR and TE were set to the shortest values available on the scanner; the recorded values ranged from 1.89 to 2.04 ms and from 0.94 to 1.02 ms, respectively (Table S1).

Table S1 lists the geometry and timing of each recording. The recorded geometry comprised 20–28 sagittal slices at 6 mm spacing, with a reported slice thickness of 12 mm. The in-plane acquisition matrix, read from the DICOM header, was 116 *×* 113 for ten participants and 116 *×* 116 for one over a 400 *×* 400 mm field of view, giving nominal in-plane sampling of 3.45 *×* 3.54 and 3.45 *×* 3.45 mm, respectively. The scanner reconstructed each slice onto a 384 *×* 384 grid for two participants and 256 *×* 256 for the other nine, with stored pixel spacing of 1.04 or 1.56 mm. Acquisitions comprised 200–800 volumes per participant. Recorded volume intervals ranged from 301 to 438 ms. For one recording with no documented interval, we used 324 ms (Table S1). Each reconstructed volume was assigned one timestamp.

In the same session, static whole-body images were acquired with a two-dimensional multislice balanced turbo field echo sequence with spectral fat suppression (bTFE SPIR): flip angle 60°, TR/TE 3.3–3.8/1.7–1.9 ms, 6 mm contiguous slices and a 300 *×* 240 acquisition matrix over a 400 mm field of view reconstructed to 512 *×* 512 (0.78 mm pixels; 224 *×* 179 over 300 mm with 0.59 mm pixels for one participant). One static stack per fetus, sagittal where available, was used for body-shape estimation.

### 3.2 Image processing and whole-body reconstruction

#### Image preprocessing and analysed recordings

Image preprocessing resampled the slice direction only, expanding the 20–28 stored slices to 77–154 by Lanczos-3 interpolation (MATLAB imresize3). The in-plane grid was not resized. Segmentation, tracking and validation used a subset of the interpolated slices sampled at a fixed interval that approximated the stored slice spacing (6.2–6.3 mm), together with the end slices of the stack (22–27 slices per fetus; Table S1). In-plane distances use the DICOM pixel spacing, and through-plane distances were derived from the interpolated slice spacing and the indices of the slices used.

Across the 11 recordings, 7,030 volumes were acquired and 6,911 were retained, spanning approximately 38 min on the analysed time axes. The 119 volumes not retained preceded the first verified keyframe of their recording, and no volumes were omitted from inside the retained intervals. Per-recording volume intervals and their provenance are listed in Table S1.

#### Segmentation and keyframe propagation

We used manually specified bounding-box prompts with SAM2 to semiautomatically segment the uterine wall and 14 fetal body parts across all slices in 3–17 selected volumes per fetus. Volumes were selected to cover vigorous movement intervals. These masks seeded slice-wise SAM2 propagation through time.[42] Propagation produced segmentations at every 20th volume and at the initial segmentation volumes. All resulting segmentations were visually inspected, and incorrect part masks were manually corrected.

We use keyframe to denote any volume with verified segmentation, whether initially segmented or generated by propagation. A manual frame specifically denotes a volume with hand-drawn segmentation and is a subset of the verified keyframes. The hand-drawn reference masks used for validation are distinguished from SAM2-assisted masks below. No intermediate keyframes were generated when initial selected volumes were separated by fewer than 20 volumes.

#### Image-guided tracking between keyframes

Between consecutive keyframes, each body part was treated as a rigid shape defined by its verified segmentations at the two bounding keyframes. The pose of each part at each intermediate volume was estimated by image-guided registration of an intensity template, initialised by forward and backward tracking from the two keyframes. The poses of all parts were then optimised jointly over the interval, with terms for image similarity, non-penetration between nonadjacent parts, connectivity at shared joints and temporal smoothness. The optimised poses transported the keyframe shapes to each intermediate volume, and their distance-weighted blend defined the output segmentation.

#### Whole-body pose reconstruction

We fitted the Skinned Multi-Infant Linear model (SMIL), a statistical infant body model, to the tracked segmentation at each frame.[28, 43] Keyframes were fitted first, and intermediate frames were fitted in forward and backward chains from the two bounding keyframes and blended.

The data term was a probability-weighted optimal-transport distance between each part’s model vertices and the corresponding segmentation surface points, regularised by body-shape, pose and joint-limit priors. Each intermediate pose was then constrained to remain near the path between the poses of its two bounding keyframes. Body shape and global scale were estimated once per fetus from the static stack, which was segmented into the uterine wall and the 14 body parts with the same procedure and fitted with the same objective, and were held fixed for all dynamic frames, which were fitted for pose and translation only.

#### Geometric and temporal postprocessing

No temporal smoothing was applied to the fitted poses before postprocessing. Rotation vectors were only re-expressed to be continuous in time (choosing, for each frame, the representation of the same rotation closest to the previous frame), which leaves the geometry unchanged. The postprocessed sequence is the single starting point of all contact, kinematic and muscle-length analyses.

We jointly postprocessed each subject’s fitted pose sequence to reduce geometric inconsistencies. The objective combined the distance to the fitted vertices weighted by part visibility, a penalty for penetration between nonadjacent parts, temporal smoothness that was stronger for poorly observed parts, and a penalty on joint angles outside the subject’s observed range.

#### Temporal interpolation and inverse kinematics

Denoising acted on the three-dimensional joint positions of the postprocessed body model at the acquisition rate, before any interpolation, using a 3-sample median filter followed by total-variation denoising (weight 0.5 mm). Joint positions were then interpolated to 24 Hz with shape-preserving piecewise cubic interpolation on the subject-specific acquisition timing,[44] and joint angles were obtained by inverse kinematics on a scaled infant musculoskeletal model in OpenSim.[29]

Frame-wise inverse kinematics introduces high-frequency jitter even from smooth markers, which was suppressed with a fourth-order zero-phase Butterworth filter at 3 Hz on the interpolated trajectories. This cutoff is a processing parameter rather than an acquisition bandwidth (native Nyquist frequency, 1.1–1.7 Hz), and it retained 83–110% of the angular speed of knee angles computed geometrically from the denoised joint positions. Pelvis orientation was filtered as a quaternion rather than as Euler angles to avoid filtering discontinuities caused by the Euler-angle singularity.

Muscle lengths, segment positions and joint-centre positions (shoulder, elbow, wrist, hip, knee and ankle) were recomputed from the filtered angles using OpenSim Analyze (MuscleAnalysis and BodyKinematics, without dynamics).[29, 45] The internal low-pass filter was disabled, and the analyses used the 24 Hz time base. These kinematic and muscle-length quantities were the input to the subsequent analyses, with the additional 1 Hz filters specified for endpoint trajectories and muscle lengths below.

The model used for inverse kinematics (HK2025 with Orig model.osim) was derived from the previously described infant-scaled Lai–Arnold 2017 model[8] and scaled to each fetus (Code availability). Inverse kinematics used OpenSim’s InverseKinematicsTool with its default convergence accuracy and per-marker weights between 0.05 and 3, as listed in the setup file of the release.

### 3.3 Movement and sensorimotor measures

#### Movement bouts

Each limb endpoint was the wrist joint centre minus the shoulder joint centre for an arm and the ankle joint centre minus the hip joint centre for a leg, expressed as a vector in the global frame in millimetres. Positions were low-pass filtered at 1 Hz (second-order zero-phase Butterworth) before differentiation. The same criteria were used for all fetuses and limbs, requiring a peak speed of at least 12 mm s*^−^*^1^, boundaries at 4.8 mm s*^−^*^1^, merging gaps shorter than 1 s, duration of at least 0.5 s and net displacement of at least 8 mm. The thresholds represent the smallest movement regarded as a bout by the authors, and surrogate trajectories built from the denoising residuals produced no bouts under them. Two fetuses had no detected bouts under these criteria.

For Fig. 4e, angular speed was the recording mean of absolute angular velocity and motion range was the 2.5–97.5 percentile range, with the sides averaged before cohort summary. For Fig. 4f, signed angular velocities were averaged in 1-s bins and Spearman correlations were calculated per fetus among the 20 limb coordinates. Coefficients were Fisher z-transformed and compared with zero across the 11 fetuses by a one-sample t-test, with Benjamini–Hochberg correction over the 190 pairs. Displayed values are the means of the Fisher-transformed coefficients across fetuses, transformed back to the correlation scale with the hyperbolic tangent.

#### Geometric estimation of skin contact

The 24 Hz skin series used for contact estimation was generated from the per-volume body-model parameters after a five-volume moving-median outlier screen (3 median absolute deviations), shape-preserving interpolation to 24 Hz, an 11-sample fourth-order Savitzky–Golay filter and a fourth-order zero-phase low-pass filter at 2 Hz applied to the pose, global orientation and translation parameters (hand poses were interpolated without smoothing); the joint series used for the kinematics was derived from the unsmoothed per-volume parameters as described above. The trajectories of all 6,890 skin vertices of this series were then processed at the acquisition times by a three-volume median and total-variation denoising (Chambolle algorithm, weight 0.5 mm), followed by shape-preserving piecewise cubic interpolation to 24 Hz and a fourth-order zero-phase Butterworth low-pass filter at 3 Hz. The median and total-variation steps displaced vertices by a mean of 0.32–0.48 mm across fetuses. Contact was recomputed from these temporally processed surfaces.

Uterine-wall proximity was evaluated per skin vertex at 24 Hz. A vertex was flagged when it lay outside the uterine-wall mesh radially scaled towards its centroid by 0.90, which defined an inner shell 9.3–12.3 mm thick across participants (median, 9.9 mm; 10% of the mean radius of the reconstructed wall mesh). Self-proximity required vertices of distinct body parts to be separated by less than 0.01 model units, corresponding to 7.3–9.1 mm (median, 7.8 mm).

Both criteria are deliberately generous relative to physical skin contact, given 6 mm slice spacing and surface residuals of 5.8–9.8 mm. The fractions therefore represent proximity rather than confirmed touch. Radial scaling makes the shell proportionally thicker where the uterus is wider. Vertex flags and their derived part-pair contact matrix were debounced with a 1.04 s (25-sample) temporal median, which changed the time-averaged fractions by at most 0.11 percentage points. Body-part-pair episodes were extracted after a further 0.5 s debounce. Pairs were retained only if their duty cycle was 5–90% with at least two episodes.

For the duration distribution in Fig. 5d and the uterine-wall episode proportions in Fig. 5e, an episode was defined at the body-part level by at least 5% of its vertices being flagged, with gaps shorter than 0.25 s merged. Episode proportions use denominators pooled across the 11 fetuses (568 body-part uterine-wall episodes for Fig. 5e and 1,499 retained part-pair self-contact episodes for Fig. 5f, each undirected pair counted once). Episodes from different parts or pairs may overlap in time, and these denominators differ from the 102 merged hand-contact events.

The hand-occupancy analysis pooled sampled frames from both hands of all 11 fetuses. Self-contact was scored by a body-part-pair flag with the head or arm segments (upper arm, forearm and opposite hand) as partners, and wall contact by any flagged hand vertex. Occupancy was the number of flagged hand-frames divided by the total number of sampled hand-frames, with fetus-level fractions also summarised by their median. Episodes were contiguous flagged runs without gap merging, and the proportion attributable to episodes lasting at least 10 s used the flagged hand-frame total as its denominator.

Spatial separation of the two contact types was summarised from the same production flags used in Fig. 5a,b. For each fetus, the time-averaged proportion of samples in which each of the 6,763 labelled skin vertices met the uterine-wall criterion and the self-contact criterion was computed separately. A vertex was assigned to a contact type when that proportion exceeded a fixed occupancy threshold, and the Jaccard index was the number of vertices assigned to both types divided by the number assigned to either. The main text reports the median across the 11 fetuses at a 10% occupancy threshold (5.4%; range, 0.3–34.2%), with 5% and 20% as sensitivity settings (medians, 7.5% and 3.2%). At the 10% threshold, the observed overlap was 0.46 times the value expected if the two assignments were independent.

For hand-contact onsets, the hand–partner gap was the fifth-smallest nearest-vertex distance over the hand vertices, and for the wall the fifth-smallest radial margin to the convex hull of the uterine surface, applying the same radial ratio as the vertex criterion to the hull. A contact began when this gap reached the production criterion (self, 0.01 model units; wall, 10% of the mean hull radius) and ended when it exceeded that criterion by 5 mm. An onset required a confirmed separated state earlier in the recording, contact present at the start generated no onset, and contacts shorter than 0.67 s were discarded. Partner classes were head, other hand, arm (other forearm or upper arm, or same-side upper arm) and uterine wall, and the hand–hand gap was the smaller of the two directional gaps and generated a row for each hand. This yielded 127 rows (22 head, 34 hand, 30 arm and 41 wall) in nine fetuses. For the principal event count, each physical hand–hand contact was counted once (retaining the right-hand row), and same-hand onsets of the same broad contact type (self-contact or uterinewall contact) within 3 s were merged. The earliest onset and its associated partner were retained within each merged group, yielding 102 events. The whole-body context summaries (Fig. S6c–f) used the 86 hand-specific self-contact onset rows before this merging.

#### Muscle lengths and data-driven grouping

Muscle length refers to the model-derived length of the muscle–tendon unit. The subject-scaled model contained 162 muscle–tendon actuators representing 68 named muscles, some of which were modelled in multiple compartments.[8, 46] We refer to these actuators as modelled muscles. The inverse-kinematics solution drove their lengths at 24 Hz. Two auxiliary translations (thorax–neck and lumbar, allowed *±*10 mm during fitting) were locked at zero for all muscle-length and reference-length calculations.

A single reference posture was defined for the cohort. For each fetus, local maxima of summed endpoint speed over the four limbs, of at least four times the bout boundary speed (19.2 mm s*^−^*^1^) and at least 0.5 s apart, were identified. Joint coordinates at those times were averaged within each fetus, then across the 11 fetuses with equal fetal weighting. Left and right were pooled. Lumbar extension was retained, while lumbar bending and rotation and cervical coordinates were set to zero. The resulting posture had hip flexion 107°, knee flexion 112°, elbow flexion 94°, shoulder elevation 32°, lower lumbar extension *−*22° and lumbar extension *−*8°. Each fetus’s scaled model in this posture supplied the reference length *L*_ref_*_,m_* for every muscle. This posture is a common denominator for normalisation and is not a physiological resting or optimal length. The complete reference coordinates are included in Source Data.

Length deviation was 100[*L_m_*(*t*) *− L*_ref_*_,m_*]*/L*_ref_*_,m_*. Each normalised length series was low-pass filtered at 1 Hz (second-order zero-phase Butterworth) before excursion analysis or differentiation.

The signed rate was *q_m_*(*t*) = 100[*dL_m_*(*t*)*/dt*]*/L*_ref_*_,m_*, in % of reference length s*^−^*^1^. Positive lengthening rate was the temporal mean of max(*q_m_*(*t*), 0), with zero contribution while shortening. Absolute rates *|q_m_*(*t*)*|* were used for regional summaries, whereas signed rates were used for muscle covariation and event comparisons.

Fig. S5 reports the recording excursion of each muscle (maximum minus minimum of the filtered series, in % of *L*_ref_*_,m_*), averaged within eight anatomical regions (neck, 8 muscles; trunk, 26; shoulder, 18; upper arm, 14; forearm, 20; hip, 38; thigh, 16; calf, 22).

The main-text excursion summary is the median across the 162 muscles within each fetus and then across the 11 fetuses, with the range describing the fetus-specific medians. For the module-peer summary, each assigned muscle’s signed rate was regressed on the unweighted mean of the other muscles’ signed rates in its module using 1-s bins, and the coefficients of determination were summarised by the median across muscles within each fetus and then across fetuses.

To relate muscle lengths to the limb-movement bouts defined above, each of the 128 limb muscles was assigned to the limb containing its path (shoulder, upper-arm and forearm muscles to the arm of the same side, 26 per side; hip, thigh and calf muscles to the leg of the same side, 38 per side). For each limb, the positive lengthening rate was averaged over the frames inside the bouts of that limb and over the remaining frames. The 34 neck and trunk muscles were not assigned to a limb and were averaged over the frames in which any limb was in a bout. Limbs without a detected bout were excluded, leaving 31 limbs in nine fetuses, and fetuses 2 and 4 contributed no bout at all. Within each fetus, the samples of its eligible limbs were pooled before averaging, and the within-bout and outside-bout means were compared across fetuses with a two-sided Wilcoxon signed-rank test (Fig. S7). Figures 6c and 6d use the within-bout averages, whereas Figs 6e and 6f use the full recordings of all 11 fetuses.

For Fig. 6e, the signed rate of each muscle was averaged in 1-s bins. Within each fetus, pairwise Spearman coefficients were calculated, Fisher z-transformed, averaged across the 11 fetuses and transformed back to the correlation scale. For Fig. 6f, module-mean signed rates (unweighted means of the member muscles, including one unassigned muscle as a separate row) were similarly averaged in 1-s bins and correlated by Spearman within fetus. The displayed off-diagonal coefficients are means of the Fisher-transformed coefficients across fetuses, transformed back to the correlation scale, with diagonal cells hidden, and module pairs were tested against zero across fetuses by a one-sample t-test on the Fisher-transformed coefficients with Benjamini–Hochberg correction over the 528 pairs. For Fig. 6d, the rate for a module is the unweighted mean across its member muscles of each muscle’s mean positive rate within bouts. Thus the positive-part operation precedes averaging across muscles.

Modules were derived from the signed rates without anatomical labels, treating all 162 right- and left-side muscles as distinct series in a joint analysis. Each recording was divided into 5-s windows with a 2.5-s step. Within each window, muscle-rate series were linearly detrended and multiplied by a Hann window. Pearson muscle-pair correlations were Fisher-transformed, averaged over windows within each fetus and then equally across fetuses, and transformed back with the hyperbolic tangent. Affinity propagation[47] used the resulting precomputed similarity matrix (off-diagonal median as preference; damping 0.9; maximum 2,000 iterations; convergence window 30 iterations; random state 0). The number of groups was not specified at this stage.

Stability was assessed with 1,000 resamples. In each, 11 fetuses were sampled with replacement and windows were resampled within each selected fetus. The co-assignment matrix *A* recorded the fraction of resamples assigning a pair of muscles to the same group. Bootstrap group counts had median 33.5 (interquartile range, 32–35; range, 26–38), and the average-linkage consensus of 1 *− A* was cut at K = 33, giving 32 modules of 2–11 muscles and a singleton (R SUP) left unassigned. For each assigned muscle, the stability score was its mean co-assignment with the other members of its module (excluding itself). Module scores were the means of their member scores. The 0.8 threshold was used to summarise stability and to shade the 14 modules with mean scores below this value in Fig. 6c.

Module series are unweighted means of member muscles. Modules are ordered as axial or bilateral groups, right arm, left arm, right leg and left leg. Within the left-side blocks, matched modules follow the order of their right-side counterparts according to mirrored membership (Table S2). Tentative names use the most frequent functional label obtained by regressing muscle length on joint coordinates. Groups with a reported purity below 0.5 are labelled mixed. Sensitivity runs varied window length, resampling scheme (whole-recording or moving-block), fetus subsampling and mirroring, and were compared with the production partition by label-invariant adjusted Rand indices (Table S3).

### 3.4 Multimodal and hand-contact analyses

#### Regional multimodal series

Skin vertices were assigned to six regions (head, trunk, right arm, left arm, right leg and left leg), excluding 127 unlabelled vertices and retaining 6,763, and the 162 muscles to six corresponding groups, comprising neck (8 muscles), trunk (10), right and left arm (32 each) and right and left leg (40 each). Head skin was paired with neck muscles. At each 24 Hz sample, the uterine-wall and self-proximity fractions of a region were the percentages of its labelled vertices carrying the respective production flag. The two flags were not treated as mutually exclusive.

Signed muscle rates were *q_m_*(*t*) = 100[*dL_m_*(*t*)*/dt*]*/L*_ref_*_,m_*, using the same reference posture and 1 Hz length filtering as above, and regional rates were unweighted means of *|q_m_*(*t*)*|* over the muscles of each group. Rates of change of contact were the time derivatives of the contact fractions after a 0.33 s moving average (Fig. 7b,c,f), or the first differences between consecutive 1-s bins (Fig. S8). Because the contact fractions are percentages of regional vertices, their rates of change are expressed in percentage points per second, whereas muscle-length-change rates are expressed as a percentage of reference length per second. For the whole-recording analyses, series were averaged in non-overlapping 1-s bins, excluding the first and last 2 s, which left 59–269 valid bins per fetus. In Fig. 7d, each series was first averaged over the valid bins of each fetus and then summarised across the 11 fetuses. Fig. 7e pools 2,252 valid 1-s bins from all fetuses in 20 *×* 20 histograms of contact extent (the sum of the two fractions, clipped at 100%, which approximates their union) against the regional muscle rate, and is descriptive only.

For the whole-recording correlation matrix (Fig. S8), the 18 regional series (12 rates of change of contact and six muscle-group rates) were rank-transformed over each fetus’s valid bins. For each pair, both series were regressed on the ranked mean rate of the muscle groups not in the pair, and the Pearson correlation of the residuals gave a pair-specific partial coefficient. This conditions each pair on a summary of muscle-length change in the remaining groups, rather than on all other variables jointly. The matrix shows cell-wise medians across the 11 fetuses. For inference, each coefficient was Fisher z-transformed and compared with zero across fetuses by a one-sample t-test, with Benjamini–Hochberg correction[48] over the 153 pairs. The same-region uterine-wall–self summary was the mean of the six regional coefficients within each fetus.

For Fig. 7f, each limb-movement bout (defined above) gave a 10 s window centred on the bout. Within the window, the rate of change of the contact fraction on each of the eight parts of the moving side (head, trunk, upper arm, forearm, hand, thigh, calf and foot) was correlated (Spearman) with the signed length-change rate of each of the 19 muscle modules on that side, bilateral modules being represented by the module of the moving side. For display, available bout coefficients from the 209 bouts in nine fetuses were averaged in Fisher z space and transformed back to the correlation scale. For inference, each cell required finite coefficients from at least ten bouts. Within each cell, we averaged the Fisher z values for each fetus contributing at least three such bouts and tested those means against zero with a one-sample t-test when at least six fetuses qualified. Thus 298 cells included eight fetuses, four included seven and two included six. Benjamini–Hochberg correction was applied across the 304 cells. The merged-contact analysis (Fig. S9) used the fraction of vertices carrying either flag. All 152 cells included eight fetuses and formed a separate correction family.

#### Hand-contact analyses

For the gap-closure decomposition, each onset defined a reference contact point at the hand’s position, which was carried backwards with the contacted segment (skull, other hand, radius or humerus) for self-contact and held fixed in the global frame for wall contact. This reference-point gap, distinct from the skin gap used for detection, was zero at onset by construction. Its closure during the last 2 s was decomposed, in the thorax frame, into hand displacement and contact-point displacement for self-contact, and into hand displacement relative to the thorax and transport of the hand by the trunk for wall contact. A contribution was dominant when its share exceeded two thirds, and closure below 3 mm was labelled unchanged. Lengths were converted to millimetres using each acquisition’s in-plane pixel size.

For the internal consistency check (Fig. S6a), closing speeds derived from skeletal joint-centre positions were aligned to the skin-based onsets and compared with control times 6–14 s away in the same recording, with up to four controls per onset and controls at least 3 s from any onset. Each physical hand–hand contact was counted once (17 contacts), as its two hand-specific onset rows describe an identical distance curve. Because this construction also favours apparent closure at control alignments, only onset–control differences were interpreted, using the mean closing speed from *−*1 to 0 s and within-fetus permutation of onset and control labels (20,000 permutations).

For the contact-family analysis (Fig. 8d), muscle-length-change rates were averaged over *−*1.5 to *−*0.4 s before each onset. Excluding the immediate 0.4 s neighbourhood does not provide sub-volume temporal resolution. All 102 counted onsets had at least one local control window 6–14 s away. Seven fetuses contributed uterine-wall contacts (41 onsets), four contributed head contacts (21) and seven contributed arm contacts (40, pooling contacts with the opposite hand, the opposite forearm and the same-side upper arm).

The 17 test quantities comprised six regional rate magnitudes defined relative to the touching side (touching arm, other arm, neck, trunk, same-side leg and other leg), one whole-body summary and the signed rates of the ten modules of the touching arm (seven) and neck (three), referred to by their predominant actions (Table S2). For regional magnitudes, we first averaged signed rates over the muscles in each module at each time point, took the absolute value and then averaged over the window. Regional values were unweighted means of these module magnitudes. For the ten individual module measures, we averaged the signed module rates over each window. For each measure, the mean of the local control windows was subtracted from the preonset value. The whole-body summary was the unweighted mean of the six regional summaries. Thus, these event-analysis magnitudes use absolute module-mean rates, whereas the regional summaries in Fig. 7 average the absolute rates of individual muscles.

Within each contact family, the 17 quantities formed one Benjamini–Hochberg correction family. The test statistic was the mean onset–control difference over onsets, so that fetuses with more onsets carried more weight. The null distribution was obtained by permuting, within each onset’s set of windows, which window was labelled the onset (10,000 permutations). Bars in Fig. 8d show this event-weighted mean, and error bars show the s.e.m. of the fetus means of the same differences. The same windows, controls and tests were applied to all 102 onsets pooled, with a separate correction over the 17 pooled tests. These tests estimate onset–control differences within each family. Direction agreement was summarised among fetuses with at least two onsets. Onsets and controls can overlap within a recording, which limits event-level inference.

For direct comparisons between families, onset–control differences were averaged within each fetus and family, and the difference between two families was averaged over the fetuses that contributed at least one onset to each. Significance was assessed by permuting the family labels among the events of each fetus and recomputing the fetus-averaged statistic (10,000 permutations), with Benjamini–Hochberg correction over the same 17 quantities within each pairwise contrast. Thus, fetuses supplied the means and events within fetuses supplied the labels for permutation. Robustness to individual fetuses was assessed by repeating the family analysis with each fetus left out in turn (5,000 permutations per run) and recording whether the sign and the within-family *q* of each effect were retained (Table S4).

For the whole-body context, co-occurring contacts were counted during the first second of hand self-contact, and the number of other limbs in a detected movement bout was counted during the 2 s before onset and at local controls (Fig. S6c,d,f). Summed absolute normalised muscle-length changes over the same preonset interval were partitioned by body region (Fig. S6e). These whole-body context summaries are descriptive.

### 3.5 Validation and sensitivity analyses

#### Validation protocols

We evaluated segmentation in three protocols using three-dimensional Dice scores against hand-drawn reference masks. Joint-configuration agreement was assessed in a fourth protocol using landmarks created specifically for validation after reconstruction had been completed.

(i) Keyframe segmentation. In a leave-one-manual-frame-out evaluation (138 cases, 8 fetuses), each target manual frame was predicted from two other hand-drawn segmentations of the same fetus using sparse SAM2 propagation conditioned on both. The three configurations were interpolation, forward extrapolation and backward extrapolation. The held-out frame was removed from both the manual-frame and propagated-keyframe inputs, and propagation was rerun without its segmentation mask.
(ii) Keyframe segmentation at controlled spacing. Targets were manual frames (185 cases, 10 fetuses), and the reference keyframes were the nearest production keyframes at least 20, 50 or 100 frames away on each side. The production keyframes were conditioned on all manual frames in the processing range, including the target. This protocol evaluated performance at controlled keyframe distances within that pipeline. Results were stratified by keyframe distance, part speed (centroid displacement per frame from the nearest keyframe) and appearance change (mean absolute intensity difference within the target annotation, as a fraction of the full intensity scale).
(iii) Articulated interpolation. One manual frame was removed and reconstructed from its neighbouring keyframes at production spacing, with and without the image term. This leave-one-manual-frame-out comparison evaluated the contribution of image guidance.
(iv) Pose-level agreement. After the reconstructions were completed, one author annotated the shoulder, elbow, wrist, hip, knee and ankle of both sides on two acquired volumes from each of the 11 study fetuses without viewing the reconstruction. Each joint was entered as an in-plane disc (centre and radius) together with the first and last acquired slices in which the joint could plausibly lie, explicitly representing in-plane and through-plane annotation uncertainty. The 22 volumes comprised 15 production keyframes and 7 intermediate volumes. Five intermediate volumes were selected within the fastest movement window of that fetus and two within a quiet interval. One elbow could not be identified, yielding 87 elbow- or knee–volume pairs.

The primary quantity was the three-point configuration angle subtended at the elbow or knee, computed identically from the annotated centres and the corresponding reconstructed points. For each annotated triplet, 4,000 samples were drawn uniformly within the three discs and slice ranges to propagate annotation uncertainty to an angle distribution. Agreement was summarised with Lin’s concordance correlation coefficient[49], Spearman correlation, signed bias and interval coverage. For calibration, we compared the quantiles of reconstructed angles within their annotation distributions with a uniform distribution using the one-sample Kolmogorov–Smirnov test (Fig. S2c). Because joint–volume pairs are nested within 11 fetuses, limits calculated by treating pairs as independent are descriptive rather than population confidence intervals. This angle is distinct from the inverse-kinematics flexion coordinate. No reference joint label was used for fitting or tuning.

Fit residuals were computed as the median distance between fitted skin vertices and the segmented boundary. Penetration was quantified as the maximum capsule overlap between nonadjacent parts per frame. As an external reference, the whole-fetus segmentation network distributed with SVRTK[23] was applied to the same manually annotated frames (Fig. S1d).

#### Sensitivity analyses

To assess the sensitivity of the downstream estimates to pose errors informed by the landmark comparison (Fig. S10), the reconstructions of fetuses 7 and 10 were perturbed in three ways. A constant offset reduced elbow and knee flexion by 10.6°, the mean bias against the landmark annotations. Random errors with a standard deviation of 8.5° were added independently to shoulder elevation, elbow flexion, hip flexion and knee flexion on both sides, either varying slowly (band-limited below 0.1 Hz) or more rapidly (band-limited below 1 Hz), the faster-error scenario. Under an additive, independent-error approximation, this magnitude is a conservative rounding of the spread that remains when the annotation uncertainty (10.1°; Fig. S2) is removed in quadrature from the limits of agreement of the annotation comparison (25.6°, corresponding to a standard deviation of 13.1°). Combined offset and random errors were also applied. For the contact estimates, the distal segments of the skin mesh were rotated about the corresponding joint centres by these angles, ignoring the image evidence and the uterine boundary that constrain the actual fit, and the production contact criteria were recomputed on every fourth frame. For the muscle estimates, the same angles were added to the inverse-kinematics coordinates and the muscle analysis was repeated. Perturbed and unperturbed series were compared by the correlation of regional contact fractions, of per-muscle and regional length-change rates in 1-s bins and of module rates, by the change in time-averaged contact fractions and by the ratio of mean absolute rates. To extend the comparison to the analyses of Sections 1.6 and 1.7, the bout-centred correlation matrices of the two fetuses (27 and 50 bouts) were recomputed from the perturbed contact fractions and muscle lengths and correlated cell-wise with the unperturbed matrices, and the preonset responses of their 38 counted hand-contact onsets were recomputed from the perturbed muscle lengths with the same onset and control windows and compared with the unperturbed responses by correlation and sign agreement across event–measure pairs.

Contact-threshold sensitivity was evaluated on the same processed reconstructions using criteria of 2–16 mm (Fig. S4). The figure separates occupancy and episode summaries without onset re-detection (panels a–c; episode boundaries can vary with the criterion) from threshold-specific onset re-detection (panels d–f). The sensitivity analysis used the convex-hull wall shell of the onset detector (9.9–13.6 mm), and event-set overlap was evaluated within *±*1 s of the reference detections. At each of the eight thresholds from 2 to 16 mm, Spearman correlations compared fetus-level contact extent with the primary-analysis values. Across thresholds, these correlations had medians of 0.92 for self-contact (range, 0.84–0.95) and 0.98 for uterine-wall contact (0.95–0.99). These analyses assess geometric definitions, not accuracy relative to observed physical touch.

### 3.6 Implementation and reporting

#### Implementation and availability

Segmentation and tracking used Python 3.10.19 and PyTorch 2.5.1 (CUDA 12.4). Segmentation used SAM2.1 (commit 2b90b9f, checkpoint sam2.1 hiera large; configuration sam2.1 hiera l). Fitting used Python 3.8.20, PyTorch 2.4.1 (CUDA 12.1) and the neutral SMIL model with an eight-component Gaussian-mixture pose prior. Joint-centre, inverse-kinematics and muscle analyses used OpenSim 4.6, with Python 3.11.15 for the OpenSim API. Contact estimation used MATLAB R2024b Update 4, and statistical analyses used Python 3.12.2. Computation used a local workstation (NVIDIA RTX 5000 Ada, 32 GB) and rented cloud instances (NVIDIA A6000 and RTX 3090). All statistical tests were two-sided. Permutation *P* values were calculated as (1 + *b*)*/*(1 + *B*), where *B* is the number of permutations and *b* is the number for which the absolute permuted statistic was at least as large as the absolute observed statistic. All resampling and permutation procedures used fixed random seeds.

## AI-assisted preparation

ChatGPT was used for English language editing, and Claude Code was used to assist with programming.

## Supplementary information

Supplementary Movie S13 accompanies this preprint. The other supplementary figures, tables, methods and movies cited in the text (Figs S1–S10, Tables S1–S4, Movies S1–S12, S14 and S15) and further implementation details of the reconstruction are not included in this preprint and will be made available with the peer-reviewed version.

## Acknowledgements

This work was supported by JST PRESTO (grant number JPMJPR23S4 to H.K.) and the JSPS KAKENHI Grant-in-Aid for Scientific Research (S) (grant number JP25K24741 to Y.K.). We thank the participants and the staff of Kyushu University Hospital, as well as Kousei Ishigami, for their support with the imaging study.

## Author contributions

H.K. designed the study, performed the analyses and wrote the manuscript. T.W., K.S., C.T., K.N. and Y.Sa. developed the imaging protocol and acquired the MRI data. S.M., Y.Sh. and H.T. recruited the participants and provided clinical information. Y.K. supervised the project, acquired funding and revised the paper. H.K. and T.W. contributed equally to this work. All of the authors discussed the results and approved the final version of the paper.

## Competing interests

H.K., T.W., S.M., Y.Sh., K.S., C.T., K.N., Y.Sa. and Y.K. are named inventors and co-applicants on a pending Japanese patent application (no. 2025-103002, filed 19 June 2025) relating to the image-based estimation of fetal posture, joint angles, skin-surface positions and muscle-length changes described in this study. The authors declare no other competing interests.

## Data availability

The data underlying this study are not publicly available at the preprint stage. Data availability will be specified in the peer-reviewed publication.

## Code availability

The analysis code (with software environment specifications, parameters and examples of processed data) will be archived in a public repository at publication. The infant musculoskeletal model used here (OpenSim model file with the muscle paths, joint definitions and scaling settings, without display geometry) will be made available on reasonable request after publication. The bone surface meshes used for display were obtained under a commercial licence and cannot be redistributed.

## Notes

### Competing Interest Statement

The authors have declared no competing interest.

### Author Declarations

The Institutional Review Board of Kyushu University gave ethical approval for this work (approval no. 24014-00, 18 April 2024). The University of Tokyo participated as a collaborating institution under the same central review. Written informed consent was obtained from each participant.

